# The Validation of Non-Invasive Pressure-Volume Loop Indices in Severe Aortic Stenosis

**DOI:** 10.1101/2024.01.23.24301702

**Authors:** Omar Aldalati, Mehdi Eskandari, Montasir H Ali, Rita Cabaco, Jonathan Byrne, Mark Monaghan, Bobit Lukban, Nicola Kennedy, Ajay Shah, Rafal Dworakowski, Philip MacCarthy

## Abstract

**Background:** Studies utilizing invasive pressure-volume loops offer valuable insights into left ventricular (LV) contractility, yet their availability remains limited. Conversely, non-invasive indices are accessible and reproducible; however, their validation in aortic stenosis (AS) is lacking. We sought to validate the non-invasive indices of PVL studies in a group of symptomatic severe AS.

**Methods:** We recruited patients with symptomatic severe AS admitted for trans-catheter aortic valve replacement (TAVR) to undergo invasive PVL studies. Non-invasive PVL indices were measured with three-dimensional (3D) echocardiography with a pre-specified protocol. The agreement between invasive and non-invasive calculation methods were assessed.

**Results:** Eleven patients (11) were recruited for this pilot study. The non-invasive end-systolic pressure-volume relationship (ESPVR) Kelly’s method (Ees_(sb)_ = 0.9 × systolic blood pressure/end-systolic volume (ESV)) had the best agreement with invasive ESPVR (limits of agreement - 1.7 to 2.1 with a percentage error of 24%, one sample T-test p =0.504). Systolic blood pressure, as measured by the brachial blood pressure cuff, had the best agreement with end-systolic pressure in severe aortic stenosis (limits of agreement −60 to 60 with a percentage error of 3%, one sample T-test p =0.959).

**Conclusion:** Measurement of the single-beat estimate of ventricular elastance (Ees_(sb)_) is possible in patients with severe aortic stenosis. Kelly’s method (Ees_(sb)_ = 0.9 × SBP /ESV) has the best agreement with the invasive measurement of left ventricular elastance (Ees). Systolic blood pressure, as measured by the brachial blood pressure cuff, has the best agreement with end-systolic pressure in severe aortic stenosis.

## Introduction

The growing interest in early intervention for severe aortic stenosis (AS) is evident. Left ventricular (LV) ejection fraction (EF) has been used for assessment of LV function. The assessment of LV EF, regardless of the modality used, has inherent limitations. Advanced tools of LV mechanics’ assessment, such as myocardial deformation, has been developed for early detection of sub-clinical LV dysfunction.

Pressure-volume loop (PVL) indices, mainly left ventricular elastance (Ees; also known as end-systolic pressure-volume relationship (ESPVR)), are considered the gold standard measures of LV systolic function^3^.

Invasive PV loop measurement requires a left ventricular conductance catheter placement, an inferior vena cava (IVC) balloon occlusion for load variation, and a pulmonary artery catheter for calibration. The invasive nature of such a procedure renders it a research tool rather than a day-to-day investigation^4^. As a result, several single-beat non-invasive echocardiography-based methods were developed and validated for measuring Ees in clinical practice – the majority of these methods were derived from animal-based research^9^.

To the authors’ knowledge, none of the validated single-beat non-invasive methods included patients with severe AS. In this pilot study, we sought to assess the agreement between invasive and non-invasive PVL indices in severe AS patients. We also sought to compare the methods of single-beat estimates of Ees and the agreement between the other invasive and non-invasive indices of contractility.

## Methods

We recruited eleven (11) consecutive patients for invasive PV loop studies during trans-catheter aortic valve replacement (TAVR) procedures. The patients had symptomatic severe AS and were deemed by the heart team appropriate and in need of TAVR. Recruited patients had to meet the following inclusion criteria:

- In sinus rhythm
- Suitable for transfemoral access
- In a stable clinical condition
- Absence of co-existing another severe valvular heart disease
- Normal right ventricular function
- Able to give informed consent

### Invasive pressure-volume loop studies

The TAVR procedures were performed as per routine clinical care. After crossing the aortic valve, a 4 Fr or 7 Fr high-fidelity conductance catheter (CD Leycom, Netherlands) was placed in the LV, either over the guidewire or with the help of a destination catheter. The conductance catheter was then connected to a dual-field conductance processor (Inca, CD Leycom, Netherlands). Under fluoroscopic guidance, the conductance catheter position was optimised along the long LV axis and with the help of live PV loop recording.

For calibration purposes, a pulmonary artery catheter (Swan – Ganz catheter, 6 Fr, Edwards Lifesciences Corp, Irvine, USA) was placed in the pulmonary artery under fluoroscopy guidance from the right median cubital vein. Calibration was performed using the parallel conductance method. The cardiac output was measured first with thermodilution technique (SV calibration; repeated three times) followed by injection of 10 ml of 10% saline into the pulmonary artery (EFcal; repeated twice).

After calibration, we waited for several minutes to ensure cardiovascular stability. The conductance catheter position was reassessed by fluoroscopy once again before acquiring live PV loop data.

### Three-dimensional transthoracic echocardiography

All recruited patients underwent two and three-dimensional transthoracic echocardiography (2D and 3D TTE) approximately 90 minutes before the TAVR (Epic CVx, Best, Netherland). 3D LV end systolic and end-diastolic volumes (Qlab version 12, Philips, Best, Netherlands), pre-ejection period (PEP), ejection time (ET) and total systolic time (TST) were calculated. Brachial blood pressure was recorded at the time of the 3D TTE study for all patients.

### Formulas used to calculate the non-invasive Ees

The acquired TTE data was used to calculate the non-invasive Ees_(sb)_ using the following formulas:

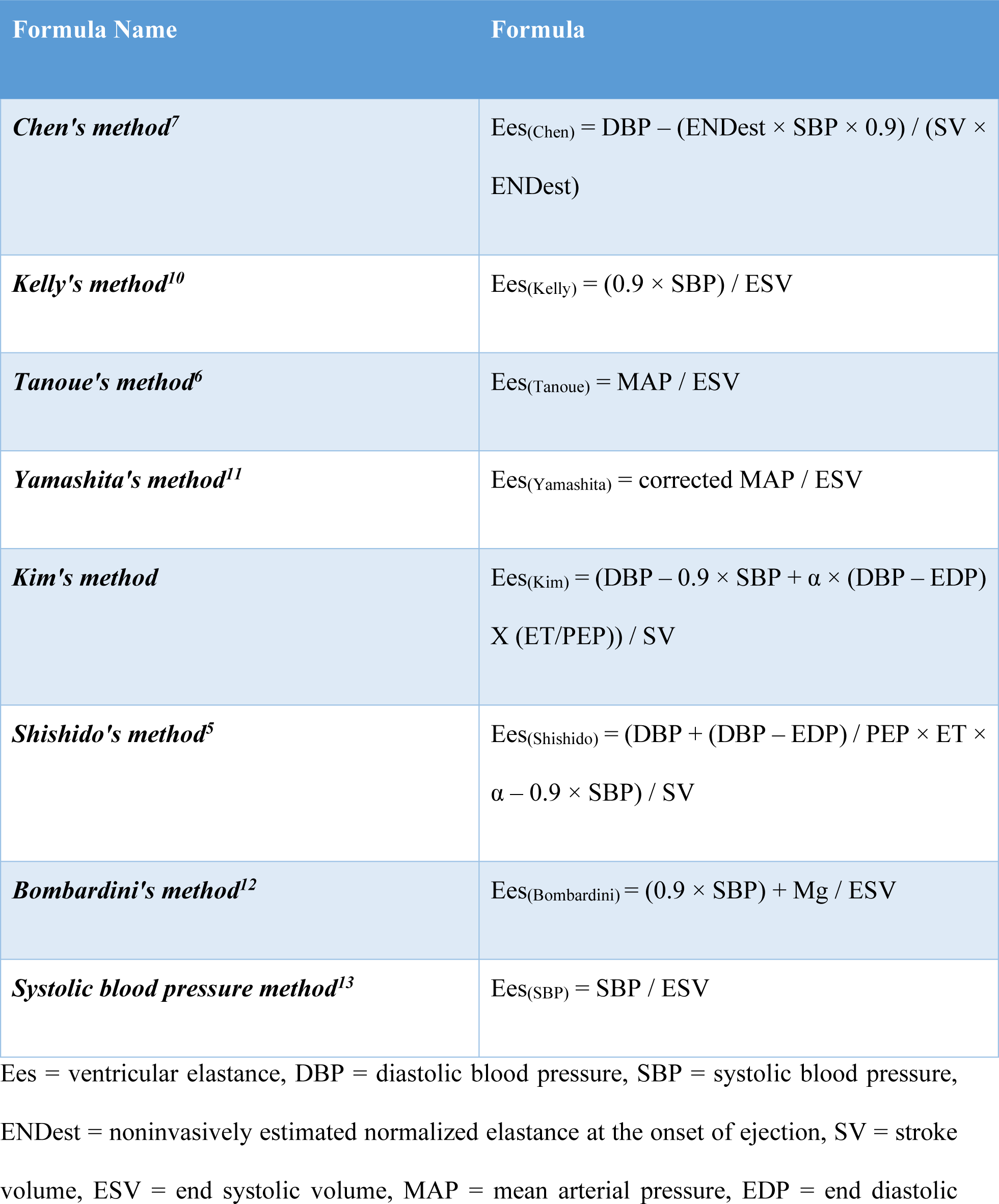
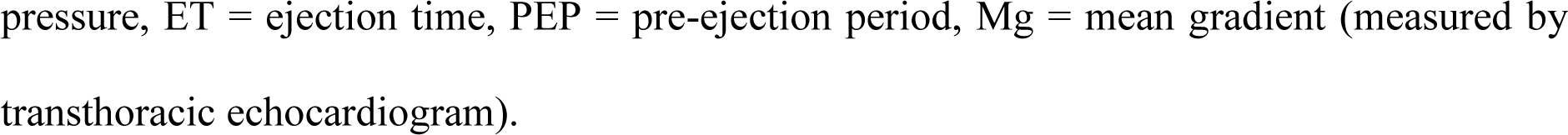

| Formula Name | Formula |
| --- | --- |
| *Chen's method*<sup>7</sup> | $E_{es(Chen)} = DBP - (ENDest \times SBP \times 0.9) / (SV \times ENDest)$ |
| *Kelly's method*<sup>10</sup> | $E_{es(Kelly)} = (0.9 \times SBP) / ESV$ |
| *Tanoue's method*<sup>6</sup> | $E_{es(Tanoue)} = MAP / ESV$ |
| *Yamashita's method*<sup>11</sup> | $E_{es(Yamashita)} = \text{corrected MAP} / ESV$ |
| *Kim's method* | $E_{es(Kim)} = (DBP - 0.9 \times SBP + \alpha \times (DBP - EDP) \times (ET/PEP)) / SV$ |
| *Shishido's method*<sup>5</sup> | $E_{es(Shishido)} = (DBP + (DBP - EDP) / PEP \times ET \times \alpha - 0.9 \times SBP) / SV$ |
| *Bombardini's method*<sup>12</sup> | $E_{es(Bombardini)} = (0.9 \times SBP) + Mg / ESV$ |
| *Systolic blood pressure method*<sup>13</sup> | $E_{es(SBP)} = SBP / ESV$ |

$E_{es}$ = ventricular elastance, DBP = diastolic blood pressure, SBP = systolic blood pressure, ENDest = noninvasively estimated normalized elastance at the onset of ejection, SV = stroke volume, ESV = end systolic volume, MAP = mean arterial pressure, EDP = end diastolic
pressure, ET = ejection time, PEP = pre-ejection period, Mg = mean gradient (measured by transthoracic echocardiogram).

The sonographers and clinicians reporting the TTE were blinded to the invasive PV loop study results. Similarly, the cardiac physiologist and the clinician reporting the PV loop studies were blinded to the results of the 3D TTE studies.

### Statistical analysis

The agreement between the invasive and the non-invasive indices was tested using Bland-Altman plots, linear regression, Wilcoxon’s test, Pearson’s correlation and the percentage error between the corresponding index^14^. Statistical analysis was performed with SPSS statistical software (IBM SPSS Statistics for Windows, Version 22.0. Armonk, NY: IBM Corp).

### Ethical approval

Approvals of the original study design and subsequent amendments were all granted by London – Dulwich Research Ethics Committee with the reference number: 13/LO/1542 IRAS project ID: 123464. All study participants gave a written consent.

The manuscript submission conforms to the guidelines set forth in the “Recommendations for the Conduct, Reporting, Editing and Publication of Scholarly Work in Medical Journals (ICMJE Recommendation)”.

### Funding

This study was supported fully by King’s College Charity and in part by a National Institute for Health Research Biomedical Research Centre award to Guy’s & St Thomas’ Hospital and King’s College London in partnership with King’s College Hospital.

## Results

Eleven consecutive patients (11) were recruited for this study. The mean age was 84 ±8 years; the majority were females (73%). All recruited patients had severe AS with a mean aortic valve area (AVA) of 0.72cm^2^ ±0.2, mean gradient of 42 ±16mmHg and dimensionless index of 0.21 ±0.06 as per table 1. The measured indices from the invasive PVL studies and TTE are summarised in table 2 and table 3, respectively.

**Table 1:** Baseline characteristics.

| Variable | AS patients (11) |
| --- | --- |
| Age (years ±SD) | 84 ±8 |
| Female | 8 (73%) |
| Body mass index (kg/m<sup>2</sup> ±SD) | 29 ±7 |
| Body surface area (m<sup>2</sup> ±SD) | 1.79 ±0.22 |
| NYHA class |  |
| II | 5 (45%) |
| III | 6 (55%) |
| Systolic blood pressure (mmHg ±SD) | 137 ±19 |
| Diastolic blood pressure (mmHg ±SD) | 73 ±12 |
| Never smoked | 9 (81%) |
| Hypertension (Yes) | 8 (73%) |
| Diabetes (Yes) | 4 (37%) |
| Peripheral vascular disease (Yes) | 3 (28%) |
| Previous myocardial infarction (Yes) | 1 (9%) |
| Previous cardiac surgery (Yes) | 1 (9%) |
| Creatinine (μmol/l ±SD) | 101 ±25 |
| Haemoglobin (mmol/l ±SD) | 124 ±9 |
| QRS duration (ms ±SD) | 84 ±18 |
| General anaesthesia (Yes) | 6 (54%) |
| **Echocardiography Characteristics** |  |
| Ejection fraction (% ±SD) | 53 ±12 |
| RWMA | 1 (9%) |
| Mean gradient (mmHg) | 42 ±16 |
| Peak gradient (mmHg) | 75 ±25 |
| Aortic valve area (cm<sup>2</sup>) | 0.72 ±0.2 |
| Dimensionless index | 0.21 ±0.06 |

AS: aortic stenosis, RWMA: regional wall motion abnormality, SD: standard deviation, NYHA: New York Heart Association

**Table 2:** Pressure-volume loop invasive measures.

| Variable | Mean ±SD | Median (IQR) | 95% Confidence Interval | |
| --- | --- | --- | --- | --- |
|  |  |  | Lower Bound | Upper Bound |
| Heart rate (bpm) | 67 ±16 | 64 (18) | 56 | 78 |
| Ejection fraction (%) | 60 ±12 | 61 (16) | 52 | 69 |
| Cardiac output (L/min) | 4.7 ±1.5 | 5 (2.7) | 3.6 | 5.8 |
| Stroke volume (ml) | 78 ±26 | 82 (56) | 59 | 97 |
| Stroke work (ml.mmHg) | 8155 ±2895 | 7766 (4089) | 6210 | 10099 |
| **Diastolic Indices** |  |  |  |  |
| End-diastolic volume (ml) | 110 ±41 | 107 (78) | 82 | 138 |
| End-diastolic pressure (mmHg) | 9.5 ±5.5 | 7.8 (10) | 5.8 | 13 |
| dp/dt minimum (mmHg/s) | -1194 ±469 | -1419 (532) | -1510 | -879 |
| TAU (ms) | 33 ±6 | 32 (12) | 29 | 38 |
| EDPVR (mmHg/ml) | 0.1008 ±0.06 | 0.1014 (0.12) | 0.0583 | 0.1432 |
| **Systolic Indices** |  |  |  |  |
| End-systolic volume (ml) | 58 ±30 | 59 (58) | 38 | 79 |
| End-systolic pressure (mmHg) | 138 ±27 | 144 (44) | 119 | 159 |
| dp/dt maximum (mmHg/s) | 1277 ±362 | 1272 (537) | 1034 | 1520 |
| ESPVR (mmHg/ml) | 3.25 ±2.1 | 2.54 (4.5) | 1.81 | 4.69 |
| SCI (mmHg/ml/s) | 14.1 ±8 | 10.5 (14) | 8.5 | 19.7 |
| PRSW (mmHG) | 82 ±52 | 67.9 (49) | 47 | 117 |
| PVA (mmHg.ml) | 12152 ±3543 | 12061 (6545) | 9771 | 14532 |
| Arterial elastance (mmHg/ml) | 2.06 ±1 | 1.74 (1.4) | 1.35 | 2.77 |
| VA coupling (Ea/ESPVR) | 0.76 ±0.33 | 0.69 (0.26) | 0.53 | 0.98 |
| ZVA invasive | 3.6 ±1.6 | 2.8 (2.7) | 2.48 | 4.74 |

bpm: beat per minute, dp/dt minimum: the minimum rate of change of ventricular pressure, EDPVR: End-diastolic pressure volume relationship, ESPVR: End-systolic pressure volume relationship, PRSW: Pre-load recruitable stroke work, PVA: pressure volume area, SCI: Starling contractility index, TAU: left ventricular time constant (diastolic index), VA Coupling: ventriculo-arterial coupling, ZVA = valvulo-arterial impedance (mmHg/ml/m2).

**Table 3:** The required non-invasive measures to calculate Ees single beat estimates, the calculated Ees and the other non-invasive measures of contractility.

| Variable | Mean ±SD | Median (IQR) | 95% Confidence Interval | |
| --- | --- | --- | --- | --- |
|  |  |  | Lower Bound | Upper Bound |
| End-diastolic volume (ml)* | 96 ±38 | 89 (88) | 70 | 122 |
| End-systolic volume (ml)* | 48 ±29 | 28 (54) | 28 | 68 |
| Stroke volume (ml)* | 48 ±14 | 44 (25) | 39 | 58 |
| Ejection fraction (%)* | 53 ±12 | 56 (15) | 45 | 61 |
| Pre-ejection period (ms) | 70 ±17 | 80 (20) | 59 | 82 |
| Ejection time (ms) | 326 ±56 | 310 (60) | 288 | 364 |
| Total systolic time (ms) | 397 ±46 | 390 (50) | 365 | 428 |
| PEP/ET (ratio) | 0.22 ±0.07 | 0.25 (0.11) | 0.17 | 0.27 |
| **Calculated Non-Invasive Ees** |  |  |  |  |
| Ees by Chen et al | 2.89 ±0.7 | 3 (0.8) | 2.4 | 3.3 |
| Ees by Kim et al | 3.1 ±1.5 | 2.8 (1.6) | 2 | 4.1 |
| Ees by Shishido et al | 11 ±4 | 9.9 (6) | 8.2 | 13.7 |
| Ees by Tanouue et al | 3.48 ±1.8 | 3.5 (3.8) | 2.25 | 4.71 |
| Ees as suggested by Bombardini et al | 4.6 ±2.2 | 5.5 (5) | 3.1 | 6.1 |
| **Other Calculated Non-Invasive Indices** |  |  |  |  |
| Arterial elastance (Ea) | 2.7 ±0.8 | 2.8 (1.4) | 2.18 | 3.31 |
| Ea as suggested by Bombardini et al | 3.6 ±1 | 4.1 (2) | 2.9 | 4.3 |
| PRSW (mmHg) | 50 ±11 | 51 (16) | 42 | 58 |
| ZVA non-invasive | 7 ±2 | 6.5 (2.7) | 5.7 | 8.4 |

\*based on 3D echocardiography.
Ea: arterial elastance, Ees: ventricular elastance (ESPVR), PEP/ET: pre-ejection period/ejection time, PRSW: Pre-load recruitable stroke work, PVA: pressure volume area, SCI: Starling contractility index, ZVA = valvulo-arterial impedance (mmHg/ml/m<sup>2</sup>).

### The agreement between invasive and non-invasive Ees (ventricular elastance)

As shown in table 4, the Chen method (considered the most reliable) was not different from invasive ventricular elastance (Ees) when compared using one-sample T-test and the Wilcoxon test. However, the correlation coefficient reached statistical significance, suggestive of a proportion bias. The Bland-Altman plot shows a clear systematic difference between the two methods. The fitted regression line suggests that for low values, Ees_(Chen)_ overestimates the invasive Ees, and the opposite is true for the higher values (Figure 1).

**Table 4:** The agreement between invasive and non-invasive indices.

| Non-Invasive Index | Mean difference to invasive index | One sample T-test | Wilcoxon's test | Pearson's Correlation | Correlation coefficient | Limits of agreement | Percentage error<sup>&</sup> |
| --- | --- | --- | --- | --- | --- | --- | --- |
| Ees by Chen | -0.359 | 0.511 | 0.722 | 0.670* | -1.136* | -3.7 to 3 | 21% |
| Ees by Kelly | 0.227 | 0.504 | 1 | 0.862* | -0.174 | -1.7 to 2.1 | 24% |
| Ees by SBP | 0.614 | 0.094 | 0.110 | 0.862* | -0.072 | -1.5 to 2.75 | 38% |
| Ees by Tanoue | -0.605 | 0.138 | 0.091 | 0.841* | -0.309 | -2.9 to 1.7 | -4% |
| Ees by Yamashita | 0.076 | 0.846 | 0.790 | 0.808* | -0.167 | -2.2 to 2.4 | 23% |
| Ees by Kim | -0.146 | 0.859 | 0.477 | 0.002 | -0.641 | -5.2 to 4.9 | 47% |
| Ees by Shishido | 7.74 | <0.001 | 0.003 | -0.050 | 1.191 | -1.5 to 16.9 | 400% |
| Ees by Bombardini | 1.358 | 0.008 | 0.003 | 0.809* | -0.641 | -1.2 to 3.9 | 71% |
| **Components of The Above Formulas** |  |  |  |  |  |  |  |
| LV ESP by Kelly | -14 | 0.153 | 0.182 | 0.160 | -0.719 | -72 to 40 | -7% |
| LV ESP as SBP | -0.5 | 0.959 | 0.929 | 0.463 | -0.050 | -60 to 60 | 3% |
| LV ESP by Bombardini | 28 | 0.008 | 0.013 | 0.296 | -0.604 | -26 to 82 | 24% |
| LV ESP as MAP | -43 | <0.001 | 0.003 | 0.207 | -0.707* | -98 to 11.8 | -29% |
| LV ESP as corrected MAP | -18 | 0.049 | 0.050 | 0.305 | -0.462 | -79 to 34.9 | -10% |
| End-diastolic volume<sup>£</sup> | -13 | 0.059 | 0.093 | 0.861* | -0.078 | -54 to 28 | -11% |
| End-systolic volume<sup>£</sup> | -10 | 0.273 | 0.350 | 0.477 | 0.135 | -68 to 48 | -11% |
| Stroke volume<sup>£</sup> | -29 | 0.001 | 0.003 | 0.710* | -0.506* | -68 to 10 | -33% |

| Other Indices |  |  |  |  |  |  |  |
| --- | --- | --- | --- | --- | --- | --- | --- |
| Ejection fraction (%) | 7.4 | 0.111 | 0.091 | 0.335 | 0.042 | -20 to 38 | 10% |
| PRSW (mmHg) | -32 | 0.037 | 0.010 | 0.742* | -1.367* | -118 to 54 | -26% |
| Arterial elastance | 0.684 | 0.025 | 0.041 | 0.611* | -0.272 | -0.9 to 2.3 | 48% |
| Ea by Yamashita | 2.66 | <0.001 | 0.003 | 0.625* | 0.282 | 0.7 to 4.6 | 150% |
| Ea by Bombardini | 1.621 | <0.001 | 0.003 | 0.632* | 0.005 | 0.1 to 3.7 | 99% |
| VA coupling* | 0.200 | 0.176 | 0.131 | -0.324 | -0.585 | -0.6 to 1 | 55% |
| VA coupling<sup>§</sup> | 0.225 | 0.199 | 0.050 | 0.346 | 0.547* | -0.8 to 2.1 | 40% |
| ZVA | 3.4 | <0.001 | 0.003 | 0.606* | 0.250 | 0.264 to 6.5 | 110% |

\*VA coupling = $Ea/Ees = (0.9 \times SBP / SV) / Ees$ (Chen), <sup>§</sup>VA coupling = $Ea/Ees = (0.9 \times SBP / SV) / (0.9 \times SBP / ESV)$ , funit = ml, &Percentage error = (mean difference / invasive reference index) × 100, Ees = ventricular elastance (mmHg/ml), SBP = systolic blood pressure, MAP = mean arterial pressure, ESP = end-systolic pressure, PRSW = pre-load recruitable stroke work, Ea = arterial elastance (mmHg/ml), VA coupling = ventriculo-arterial coupling, ZVA = valvulo-arterial impedance (mmHg/ml/m2).

**Figure 1:**
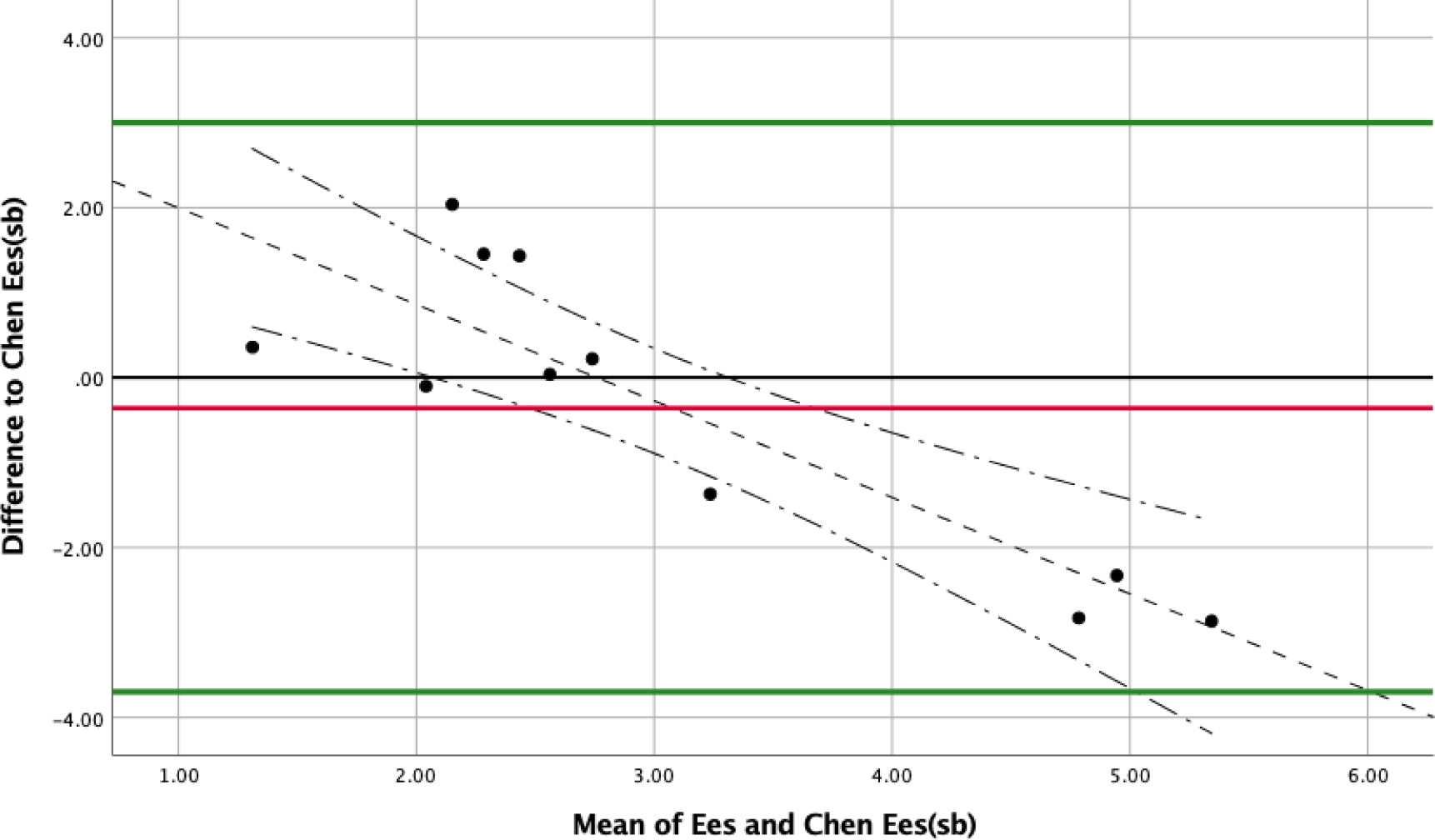
Bland-Altman plot - the agreement between Chen Ees(sb) and invasive Ees. The plot shows a clear systematic difference between the two methods. The fitted regression line suggests that for low values, Ees(Chen) overestimates the invasive Ees, and the opposite is true for the higher values.

Kelly’s method, which is by far the most widely used in non-invasive studies, has a low mean difference (0.227) that did not reach statistical significance compared to zero when tested with one sample T-test. It is also not different from the invasive Ees when compared using the Wilcoxon test. It had the highest correlation with the invasive Ees and the lowest correlation coefficient (−0.174), which did not reach a statistical difference either. The percentage error was also small (24%), with the narrowest 95% confidence interval of all methods. The Bland-Altman plot and the fitted regression line showed a homogenous data distribution around the mean excluding any systematic difference between the two methods (Figure 2). Substituting the LV ESP with SBP in the formula Ees = (ESP / ESV) ≈ (SBP / ESV) yielded similar results to Kelly’s method.

**Figure 2:**
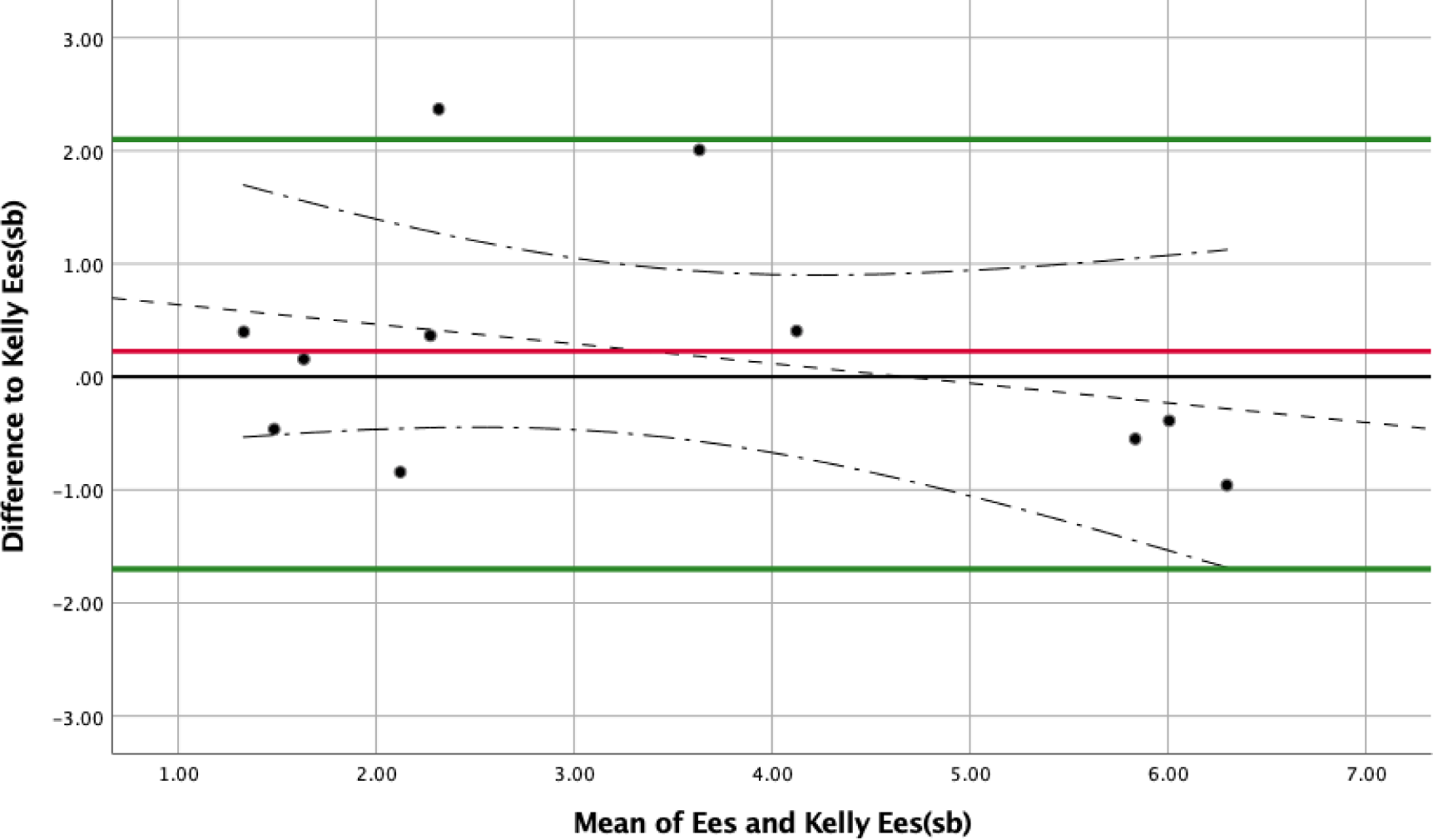
Bland-Altman plot - the agreement between Kelly Ees(sb) and invasive Ees. The plot and the fitted regression line show a homogenous data distribution around the mean excluding any systematic difference between the two methods.

The Tanoue and Yamashita methods had good agreement with the invasive Ees but were less accurate when compared to Kelly’s method as shown in table 4 (supplemental figure 1). The Kim method was not different from the invasive Ees but had a weak correlation with the invasive Ees, and the percentage error was 47%. Bland-Altman plot shows a more considerable variability compared to the above two methods (supplemental figure 2).

The Shishido and the Bombardini methods had a weak agreement with invasive Ees. The method by Shishido had the highest mean difference to invasive Ees. Both methods were statistically different when compared to the invasive Ees, and the difference reached a statistical significance when compared using the one-sample T-test. The Bland-Altman plots also show considerable variability with these methods (supplemental figure 3 and 4). The Bland-Altman plot was significant for a systematic difference when Shishido’s method was used. The Shishido method seems to overestimate the true Ees at high values.

### The agreement among the components of the single-beat estimates formulas for Ees

Using Kelly’s method, the LV ESP difference between the invasive and the non-invasive measurement did not reach statistical significance (table 4). However, the agreement between the two measures was poor as suggested by the wide confidence interval and the Bland-Altman plot (supplemental figure 5). Systolic blood pressure (SBP) had comparable confidence intervals, a smaller mean difference, correlation coefficient and percentage error to the invasive LV ESP (table 4).

The LV ESP by Bombardini, mean arterial pressure (MAP) and corrected MAP methods were statistically different when compared to the invasive ESP as shown in table 4 indicating poor agreement.

The non-invasive measures of EDV and ESV by 3D echocardiography showed good agreement with the invasive measures as per one sample T-test and Wilcoxon test. The correlation coefficients were small and did not reach statistical significance, indicative of the absence of proportional bias. The Bland-Altman plots also showed a reasonably homogenous data distribution around the mean. The SV difference between the two methods showed poor agreement as per table 4.

## Discussion

The quest to develop and validate an index that facilitates the surveillance of the left ventricular function and the determination of optimal time for intervention in aortic stenosis is evident in the recently published literature^15^. Lee et al. studied the subclinical ventricular deterioration in aortic stenosis (cardiac magnetic resonance study (CMR))^16^. One of the study’s rationale is the reduced sensitivity of left ventricular ejection fraction (LVEF) as a marker of myocardial damage. LVEF has inherent limitations irrespective of the method and modality employed. On the other hand, the European Society of Cardiology (ESC) recommends early intervention in patients with asymptomatic severe aortic stenosis and LV EF <50%^17^. The deterioration in LVEF is a late marker and usually suggestive of advanced myocardial damage which might be irreversible in some^16^.

Pressure-volume loop indices, namely LV elastance (Ees), are considered the golden standard in LV function assessment^3^. The invasive nature of such procedures limits their clinical utility. While several non-invasive methods of single-beat estimates of Ees have been developed and utilised in clinical practice, none have been validated in aortic stenosis.^12^

The Achilles’ heel in the non-invasive assessment of Ees is twofold; the measurement of LV ESP (LV end-systolic pressure) and the measurement of V0 (the maximal LV volume at which pressure is still zero).

### Estimation of LV end-systolic pressure

One of the main challenges in aortic stenosis is the estimation of non-invasive LV ESP.^12^ LV ESP in patients with no trans-aortic valve gradient, as developed by Kelly et al, is estimated as LV ESP = 0.9 × SBP; where SBP is the brachial systolic blood pressure measured by mercury sphygmomanometer.^10,18^

Kelly et al. studied ten patients (simultaneous invasive and non-invasive studies) in an attempt to calculate arterial elastance (Ea). They showed an accurate prediction of LV ESP using correlation; they did not gauge the agreement between the two methods. They also assessed another formula: LV ESP ≈ (SBP × 2 + DBP)/3 to estimate LV ESP. Both formulas had similar accuracy at predicting LV ESP (r^2^ = 0.97 and 0.96, respectively).^18^ Researchers such as Chen et al. and Kim et al. accepted this assumption (LV ESP = 0.9 × SBP) and used it in their single-beat estimate of Ees_(sb)_.^7,8^

Tanoue et al. substituted LV ESP with MAP and showed a high correlation between invasive and non-invasive Ees on an animal model of 24 mongrel dogs.^6^ However, correlation does not always mean that there is an agreement between the two methods. Moreover, the substitution of LV ESP with MAP has not been validated in humans. Chemla et al. showed that LV ESP strongly correlates with SBP but less with MAP.^19^ As such, this particular formula (MAP / ESV) has not been widely used in non-invasive studies for measuring Ees. Bombardini et al, in their non-invasive studies, substituted LV ESP with systolic blood pressure (non-invasive Ees(sb) = SBP / ESV).^12,13^

In aortic stenosis, the above assumptions fall short due to the presence of a gradient across the stenotic aortic valve (AV), i.e. the substitution of the LV ESP with a derivative of the brachial SBP would underestimate the LV ESP, and as a result Ees(sb). Yamashita et al. (co-authored by Tanoue) recognised this flaw among patients with aortic stenosis and substituted MAP with the “corrected MAP”.^11^ The corrected MAP incorporated the AV peak gradient as measured by TTE.^11^ Yet again, this assumption has not been validated in humans or in the context of AS. On the other hand, Bombardini et al. recommended the addition of the pressure drop to the brachial systolic blood pressure to estimate LV ESP.^12^

In this study, we showed that invasive LV ESP had the best agreement with Kelly’s method LV ESP when substituted with the SBP (LV ESP = 0.9 × SBP). As a substitute, SBP had the smallest mean difference and the closest correlation coefficient to zero. The other methods (MAP, corrected MAP and the Bombardini suggestion) had a weak agreement with LV ESP, as evidenced by one sample T-test and the Wilcoxon test.

### Estimation of V0

To calculate ESPVR, V0 which is the maximal volume at which pressure is still zero (the ESPVR volume axis intercept) should be measured (estimated). It is considered constant and load independent. V0 cannot be directly measured in clinical practice, but it can be estimated once Ees (the slope of the ESPVR) is known.^7^ Assuming that Ees is linear, two points from the regression line that represents Ees will be sufficient to estimate Ees (the slope) and as hence V0.^20^ To generate these two points, researchers in the past altered the LV loading conditions with inferior vena cava (IVC) occlusion and repeated the PV loop measurements. The two measures of Ees (at normal loading conditions and reduced loading conditions) constituted the two points required to estimate Ees (the slope of change in ESPVR). However, the above assumption is not entirely correct as ESPVR is nonlinear at high contractile states and low loading conditions.^7^ In large mammals, it is typically concave.^21^ Considering that ESPVR is nonlinear under many conditions, V0 becomes load-dependent. Chen et al, in their study to develop a single-beat estimate of Ees, and Maurer et al, in an echocardiography-based non-invasive survey, reported a negative V0.^7,22^

Nonetheless, the above assumptions have been generally accepted. The generated indices of contractility were still accurate, sensitive and reproducible. Chen et al. also wrote in their study: “importantly, the behaviour of the ESPVR in the physiologic loading range defines the relevant hemodynamic responses; so Ees assessed in this range is most important”.^7^

Single-beat estimates of Ees generate a single figure of Ees. Researchers such as Shishido et al., Chen et al., and Kim et al. tried to account for this fact.^5,7,8^ The used formulas were based on time-varying elastance [E(t)] during the isovolumic contraction phase and ejection phase.^9^ As such, the need for two Ees estimates at two different loading conditions has been negated. Shishido et al. then used the following formula to estimate V0: V0_(sb)_ = end systolic volume (Ves) – end systolic pressure (Pes) / Ees_(sb)_.^5^ The simplified single-beat estimate of Ees, such as Ees_(sb)_ = 0.9 × SBP / ESV, assumes V0 = zero. In this study, to simplify the research protocol in what is otherwise lengthy and risky procedures, we measured the invasive ESPVR as Ees = ESP / ESV, i.e., we assumed V0 = zero.

### The agreement between invasive Ees and non-invasive Ees(sb)

The single-beat estimate of Ees (Ees(sb)) formulas can be divided into two groups; the group that attempts to measure V0 and assumes LV ESP = 0.9 × SBP such as (Shishido, Chen and Kim) and the second group that assumes V0 = zero but substitute LV ESP differently (Kelly, Tanoue, Yamashita and Bombardini).

Chowdhury et al. studied the agreement between invasively measured Ees and non-invasive Ees(sb) among children.^3^ Their research methodology mandated vena cava balloon occlusion. They compared four different methods of Ees(sb) estimates: Chen, Kim, Shishido and Tanoue. Of note, they calculated Tanoue Ees as “Ees(sb4) = 0.9 × SBP / ESV”. Tanoue et al., in their original publication, used MAP as a substitute to LV ESP - not 0.9 × SBP. They concluded the following: Chen’s, Shishido’s and Kim’s methods over-estimated the true Ees and only the following formula Ees(sb) = 0.9 × SBP / ESV had good agreement with invasive Ees. Of note, they excluded patients with LV outflow obstruction, including patients with severe AS.

In 2014, Yotti et al. studied 27 patients with various loading conditions (eight patients with dilated cardiomyopathy, ten normal EF patients and nine patients with end-stage liver failure). Their research methodology also mandated vena cava balloon occlusion. They concluded that the Chen’s method (r2 = −0.05, p > 0.05) failed to correlate with invasive Ees while the Kelly’s method (Ees = 0.9 × SBP / ESV) had only a poor correlation (r2 = 0.38, p < 0.05).^23^

We showed that Kelly’s method (Ees(sb) = 0.9 × SBP / ESV) and the SBP method: Ees(sb) = SBP / ESV had the best agreement with the invasive Ees (allowing for the abovementioned assumptions). The methods that fall into group one had a poor agreement with invasive Ees. Likewise, the methods that assume V0 = zero (group two) but attempted to account for the gradient across the AV also showed poor agreement compared to Kelly’s method.

It seems that a simplified formula, such as Ees(sb) = 0.9 × SBP / ESV or Ees(sb) = SBP / ESV, has the best agreement with the invasively measured Ees. The above conclusion stands true regardless of the method used to estimate the invasive Ees (with or without load variation) or the studied clinical condition, including severe AS. The number of assumptions made to assemble these complex formulas is likely the reason behind these findings.

As Chen et al. suggested, ultimately, it is the sensitivity and specificity of a specific index that would determine its clinical utility. All being equal, it is the simplest method that should be used.

## Conclusion

The measurement of the single-beat estimate of ventricular elastance (Ees(sb)) is possible in patients with severe aortic stenosis. Kelly’s method (Ees(sb) = 0.9 × SBP /ESV) has the best agreement with the invasive measurement of left ventricular elastance (ESPVR). Systolic blood pressure, as measured by the brachial blood pressure cuff, has the best agreement with end-systolic pressure in severe aortic stenosis. Further studies are warranted to evaluate the efficacy of non-invasive Ees (end-systolic elastance) as a marker of left ventricular function in predicting clinical outcomes.

## Study limitations

In many prior invasive PV loop studies, protocols involved IVC occlusion to vary the load and estimate the Ees, a method we didn’t employ in this pilot study. It’s worth noting a trend among other research labs conducting these studies to simplify the invasive PVL studies.

More importantly, every preceding study, irrespective of the approach to estimate invasive Ees, indicated that Kelly’s method (Ees = 0.9 × SBP / ESV has the best agreement with invasive Ees. This aligns our findings well and bolsters their external validity.

The discrepancy observed between invasive and non-invasive SV measurements underscores concerns regarding sample size and the placement of the conductance catheter within the left ventricle. To address this, our study meticulously optimized catheter positioning via fluoroscopy and real-time PV loop acquisition. We ensured measurement accuracy by collecting data after obtaining at least two high-quality PV loop segments.

We didn’t factor in the medications administered during the TAVR procedures, including painkillers, sedatives, intravenous fluids, and, in a subset of patients, the peripheral use of vasoconstrictors like Metaraminol bitartrate.

Finally, only a few patients had mitral regurgitation. Therefore, we were unable to estimate Starling contractility index (SCI) = dp/dt Max / EDV, non-invasively.

## Clinical perspective

The growing availability of less risky and minimally invasive treatments for severe aortic stenosis (AS) is prompting earlier consideration of intervention. One emerging approach involves assessing left ventricular (LV) function and intervening based on changes in the LV. While pressure-volume loop indices are considered the gold standard for evaluating LV function, their validation specifically in severe AS is lacking.

This study aims to validate and compare several non-invasive methods for assessing PV loop indices in aortic stenosis. Our findings emphasize Kelly’s method as exhibiting the strongest agreement with invasive LV elastance measurements. This research sets the foundation for a future large-scale non-invasive study to determine the prognostic significance of these indices in aortic stenosis.

## Data Availability

Data is available upon request

## Abbreviations

AS: Aortic stenosis
EDV: End-diastolic volume
Ees: Left ventricular elastance
Ees(sb): Single-beat estimate of left ventricular elastance
EF: Ejection fraction
ESPVR: End-systolic pressure volume relationship – also known as left ventricular elastance
ESV: End-systolic volume
LV: Left ventricle
LVEF: Left ventricular ejection fraction
LV ESP: Left ventricular end-systolic pressure
MAP: Mean arterial pressure
PVL: Pressure-volume loop
TAVR: Transcatheter aortic valve replacement
TTE: Trans-thoracic echocardiography
V0: The maximal LV volume at which pressure is still zero

